# Machine Learning Models for Dynamic Assessment of Extubation Readiness in Pediatric Critical Care

**DOI:** 10.1101/2025.02.06.25321800

**Authors:** Yuxuan Liu, Sofia Cuevas-Asturias, Emma C Alexander, Jake Ormond, Tuan Chen Aw, Angela Aramburo, Samiran Ray, A. Aldo Faisal, Padmanabhan Ramnarayan

## Abstract

**Importance:** Determining the optimal timing for extubation in critically ill children remains challenging, with premature extubation leading to increased morbidity and mortality, while prolonged ventilation exposes patients to ventilator-associated complications.

**Objective:** To develop and validate machine learning models for dynamic assessment of extubation failure risk (nowcasting) and extubation readiness (forecasting) in mechanically ventilated children.

**Design, Setting, and Participants:** Retrospective cohort study using electronic health records from two pediatric intensive care units in London, UK (2013-2022), including 3,815 ventilation episodes in children aged 0-17 years.

**Exposure(s):** Mechanical ventilation via endotracheal tube using pressure control-BIPAP (PC) or spontaneous pressure support CPAP (PS) modes.

**Main outcomes and measures:** Primary outcomes were extubation failure (requiring reintubation within 48 hours) and extubation readiness (successful extubation within 12 hours). Both models incorporated demographic, physiological, ventilation, and medication data, with varying historical context lengths to optimize prediction accuracy.

**Results:** The median age of children in the study cohort (n=3815 ventilation episodes) was 8.0 months (56.2% male); extubation failure occurred in 315/3815 (8.3%). The nowcasting model achieved an area-under-the-receiver-operating-characteristic curve (AUROC) of 0.77. The forecasting model reached an AUROC of 0.85. Ventilation parameters dominated the nowcasting model, while medication response and patient characteristics drove the forecasting model.

**Conclusions and Relevance:** Our dual-model approach offers a structured framework for extubation decision-making in critically ill children, combining continuous monitoring of readiness with snapshot assessment of extubation failure risk. Prospective validation is needed; however, this strategy may help clinicians optimize the timing of ventilation liberation within pediatric intensive care.

## Introduction

Nearly 60% of children admitted to pediatric intensive care units (PICUs) in the United Kingdom receive invasive mechanical ventilation (IMV).^1^ Although IMV is a lifesaving intervention, prolonged ventilation can result in several adverse outcomes, including ventilator-associated pneumonia (VAP), delirium and muscle atrophy secondary to prolonged sedation and muscular blockade, alongside the psychological impact of ventilation on both children and their families.^2–5^ Conversely, premature extubation may lead to extubation failure (EF), which is associated with increased morbidity, longer PICU length of stay, and mortality rates as high as 20%.^6,7^ Therefore, clinicians must carefully balance the risks of continued IMV against those of premature extubation.

Current international guidelines for ventilator liberation recommend extubating a child once clinical stability is achieved, with extubation readiness assessed through a spontaneous breathing test.^8^ However, additional factors influencing EF likelihood include the risk of upper airway obstruction, level of sedation, effectiveness of cough and ability to manage oropharyngeal secretions.^9,10^ Observational studies have identified certain risk factors that predict EF in children, such as history of pneumonia, IMV duration, lower weight, stridor, and steroid treatment prior to extubation.^6,11^ However, accurately predicting EF in individual cases remains challenging.^6,11^ The decision to extubate in children continues to rely on clinical judgement, with EF rates of 5-15%, highlighting the difficulty in accurately predicting extubation success.^12,13^

The potential of artificial intelligence (AI) in aiding complex clinical decision-making in critical care has garnered increasing attention. Machine learning (ML) models trained on extensive ICU datasets have shown promise in predicting EF in ventilated adults.^14,15^ Similar models have been published in premature newborns, albeit using smaller datasets.^16,17^ However, research in critically ill children remains limited. Given differences in age-dependent physiology and disease processes, existing models cannot be directly applied to the pediatric population. Previous studies have focused on retrospective analysis of physiological and ventilation snapshot data immediately before extubation to predict EF.^9,10^ In contrast, a joint nowcasting and forecasting^18^ decision tool for predicting extubation outcomes could guide ventilation parameter adjustments, sedation dosing, and enhance clinicians’ ability to communicate accurately with parents and caregivers.^19^

This study aimed to develop and evaluate novel ML-based models for real-time extubation readiness assessment in mechanically ventilated children using a large granular multicentre data repository.

## Methods

This study was designed and is reported in concordance with TRIPOD-AI guidelines for clinical prediction models using machine learning methods.^20^ Ethical approval was granted by the United Kingdom Health Research Authority and Research Ethics Committee (23/HRA/0416).

### Study data

In this retrospective cohort study, we included routinely collected electronic health record (EHR) data from admissions between 1st January 2013 and 31st December 2022 of children aged 0-17 years and requiring invasive ventilation at two PICUs in London, UK. One unit was a 15-bedded general medical-surgical PICU, and the other a 16-bedded pediatric cardiac/respiratory ICU. Within each admission, we identified one or more ventilation episodes. A complete ventilation episode commenced with the initiation of ventilation through an endotracheal tube and ended with a successful extubation (no reintubation within the subsequent 48 hours). Incomplete ventilation episodes, resulting from death or transfer out before or within 48 hours of extubation were excluded (since many deaths in PICU occur following compassionate extubation as part of end-of-life care),^21^ as were tracheostomy ventilation episodes. Additionally, episodes were excluded under the following conditions: short-duration ventilation (≤12 hours), prolonged ventilation (>28 days),^22^ use of ventilator modes other than pressure control (PC) or pressure support (PS) (see eTable 1 in the Supplement for detailed definitions), or where episodes had missing vital sign data in more than 30% of timestamps. Since the dataset did not have documentation of formal intubation and extubation times, we inferred ventilation status at each hour by applying clinically informed rules, cross-referencing airway status, ventilator modes, and recorded ventilator parameters. Only the first complete ventilation episode during each admission was retained for analysis.

### Data extraction, pre-processing, and features of interest

Data extracted from the EHRs included patient demographics (age, sex, weight, ethnic category, diagnostic group, severity of illness at admission using the Paediatric Index of Mortality-3 score [PIM-3 score]); vital signs, airway status and ventilator readings charted on an hourly basis; blood gas parameters; laboratory blood tests; medications (sedative and neuromuscular blockade infusions, furosemide, vasopressors, corticosteroids) and discharge outcomes (mortality, length of PICU stay). A total of 37 variables were considered features of interest, as described in eTable 1 in the Supplement.

All the considered features were examined for outliers and errors utilising Tukey’s method of frequency histograms and univariate statistical methods.^23^ Where feasible, errors were rectified, such as converting tidal volume measurements from litres to millilitres. In instances where correction was not possible, values were designated as missing for subsequent imputation. Additionally, all parameters were capped at clinically plausible limits.^8,24^ To address missing or irregularly sampled data, we employed a time-limited, parameter-specific sample-and-hold approach, which aligns with common practices in health data time series analyses and intuitively mirrors the decision-making process of clinicians.^25^ For the remaining missing data, we utilized multivariable nearest-neighbour imputation, as our predictors required complete cases.^26^

### Task Definitions

We designed two tasks to assess patient readiness for extubation (Figure 1):

1. **Nowcasting extubation outcome:** The task involves assessing the probability of extubation failure at the moment of planned endotracheal tube removal.
2. **Forecasting extubation readiness:** This task entails the ongoing evaluation of the probability of successful extubation over the next 12 hours.

**Figure 1.**
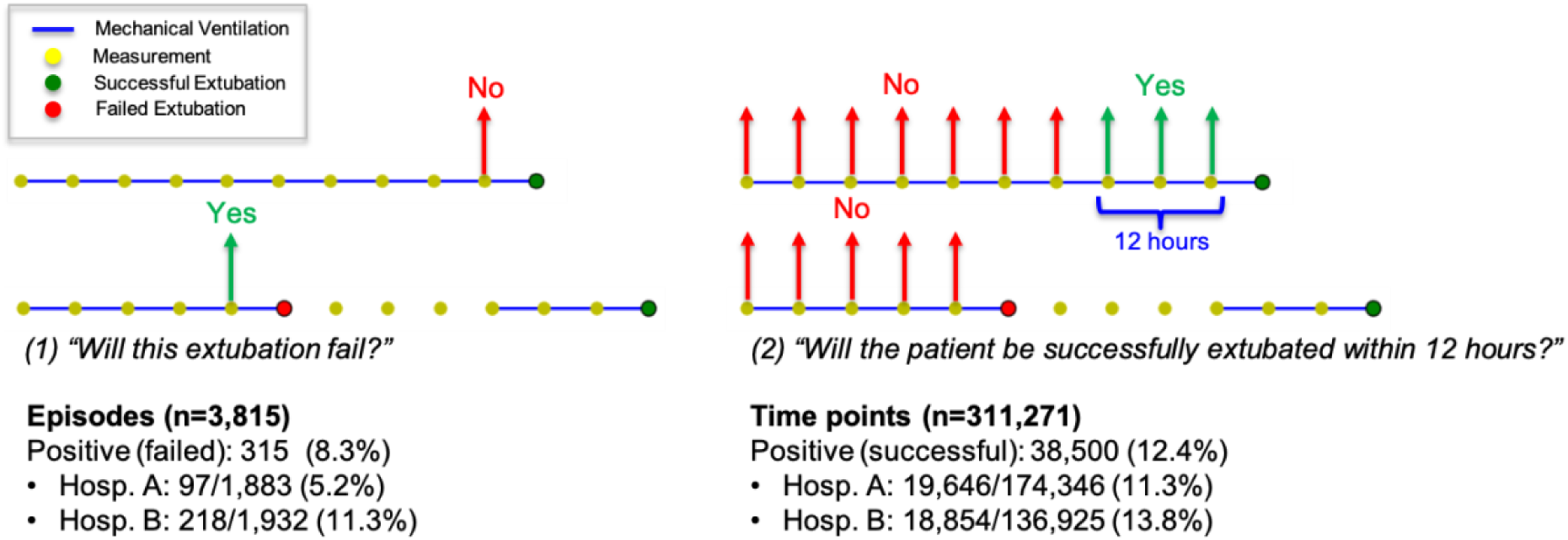
Extubation Prediction Tasks. Illustration of the two prediction tasks: nowcasting extubation failure at the time of extubation (left) and forecasting successful extubation within 12 hours (right). Blue lines indicate mechanical ventilation periods, yellow dots show measurement points, and green/red dots represent successful/failed extubations. For the nowcasting task, extubation failure rates were 5.2% (97/1,883) in Hospital A and 11.3% (218/1,932) in Hospital B. For the forecasting task, successful extubation within 12 hours occurred in 11.3% (19,646/174,346) and 13.8% (18,854/136,925) of time points in Hospitals A and B, respectively.

While extubation failure commonly occurs within the first 12-24 hours after extubation,^27^ we formally defined EF as the need for endotracheal tube reinsertion within 48 hours post-extubation, in accordance with established pediatric extubation criteria.^28–30^ This study focused exclusively on first extubation attempts, as subsequent attempts may have different risk profiles and clinical considerations. The distinct prediction targets reflect divergent clinical priorities: avoiding adverse events at the point of decision-making (Task 1) versus identifying optimal opportunities during continuous monitoring (Task 2). In both instances, the statistical convention of predicting the minority class was adhered to.

### Empirical protocols and prediction models

For both tasks, we developed predictive models focusing on the temporal analysis of patient data over time. Our methodology acknowledges the dynamic nature of a patient’s recovery trajectory and readiness for extubation, considering not only their current state, but also the progression of their condition over time. We implemented a Gated Recurrent Unit (GRU), a variant of Recurrent Neural Network (RNN), which has demonstrated superior performance in analysing sequential medical data across various clinical applications.^31^ This model’s proficiency in capturing intricate temporal patterns in patient data, makes it particularly well-suited for predicting clinical outcomes that are dependent on fluctuating patient conditions. To ensure rigorous analysis, we carried out thorough data pre-processing. Variables with normal distribution were standardized to have zero mean and unit variance, while log-normally distributed variables were log-transformed before standardization. Categorical variables were binarized and centered around zero. The distribution of each variable was assessed using visual methods such as quantile-quantile plots and frequency histograms. To determine how much historical data would optimize prediction accuracy, we evaluated model performance using different observation windows, ranging from 1 hour to 48 hours of patient data prior to each prediction time point. To handle class imbalance in our outcome data (Figure 1), we used focal loss,^32^ a method that helps the model pay extra attention to these rarer but clinically important cases.

For model evaluation, we allocated 20% of the episodes to a test dataset, with the remaining data divided into training (80%) and validation (20%) datasets. Model performance was evaluated and reported on the test dataset, using the Area Under the Receiver Operating Characteristic curve (AUROC) and the Area Under the Precision-Recall Curve (AUPRC). To ensure robustness, model evaluation was repeated five times, with different random data splits, and the average performance was reported. To provide a comprehensive assessment of extubation readiness, we integrated these two predictions into a unified risk scoring framework. The nowcasting extubation model generates a score (0-1) indicating the likelihood of failure if extubation were to occur at that moment. The forecasting extubation model produces a score (0-1) indicating the probability of successful extubation within the next 12 hours. To ensure comparability of scores, we converted the success probability to a risk metric by interpreting lower success probabilities as higher risk. This integration enables us to monitor the evolution of both immediate and prospective risks over the time approaching actual extubation events.

To understand feature contributions, we computed feature importance scores using integrated gradients,^33^ a model-agnostic attribution method that assigns importance by measuring each feature’s contribution to the model’s predictions. For sequential features processed by the GRU, importance scores were averaged as absolute values across the temporal dimension to prevent offsetting positive and negative contributions at different timepoints.^34^ The scores for each prediction task were then normalized relative to their respective maximum values to facilitate fair comparisons.

## Results

Our study included 3,815 complete ventilation episodes from 8,503 PICU admissions (detailed in eFigure 1 in the Supplement). Among the ventilated but excluded cases, data quality exclusions accounted for 768 episodes, including those with excessive missing vital signs (n=69) and instances where ventilation episodes could not be clearly classified (n=699). Incomplete clinical trajectories comprised of episodes ending in transfer to other ICUs (n=319) or mortality during ventilation (n=152). Cases excluded in line with specified exclusion criteria represented more complex clinical scenarios that necessitate a more focused investigation.

In the study cohort (n=3,815 episodes), the median age was 8.0 months (IQR: 2.0– 34.0), with cardiovascular (43.7%) and respiratory (29.5%) conditions as the predominant diagnoses, and a median IMV duration of 66.0 hours (IQR: 29.0–118.0) (Table 1). In the nowcasting model, out of all the attempted extubations, 3,500 (91.7%) were successful, while 315 (8.3%) required reintubation within 48 hours. Failed extubations occurred in younger children (median age 5.0 vs. 8.0 months, p<0.001) and were associated with significantly higher rates of neuromuscular blockade use (7.0% vs. 0.2%, p<0.001). Children experiencing extubation failure were more likely to be on PC ventilation in the hour preceding extubation rather than PS mode (65.4% vs. 35.9%, p<0.001) and required higher respiratory support as indicated by mean airway pressure (MAP: 8.0 vs. 7.0 cm H₂O, p<0.001). For the second task (forecasting extubation readiness), 42,000 (14.5%) of the 290,156 analysed time points were classified as “ready-to-extubate” based on successful extubation within the next 12 hours. At these hourly sampling points, ready-to-extubate states were associated with more favourable physiological parameters, including lower average heart rates (123 vs. 127 bpm, p<0.001) and better ventilation with lower PCO2 (5.90 vs 6.14 kPa, p<0.001) (detailed in eTable 2 in the Supplement).

**Table 1.** Demographic and Clinical Characteristics of Study Cohort.

|  |  |
| --- | --- |
|  | Overall |
| Unique ventilation episodes (N) | 3,815 |
| Age, months (Median, IQR) | 8.0 (2.0-34.0) |
| Male sex (N, %) | 2,145 (56.2%) |
| **Ethnicity (N, %)** |  |
| White or White British | 1,693 (44.4%) |
| Black or Black British | 500 (13.1%) |
| Asian or Asian British | 690 (18.1%) |
| Others (including missing) | 932 (24.4%) |
| **Primary diagnosis (N, %)** |  |
| Cardiovascular | 1,667 (43.7%) |
| Respiratory | 1,124 (29.5%) |
| Neurological | 406 (10.6%) |
| Gastrointestinal | 56 (1.5%) |
| Infection | 160 (4.2%) |
| Others | 402 (10.5%) |
| PIM-3 predicted probability of death (Median, IQR) | 0.020 (0.008-0.053) |
| Admitted with Mechanical Ventilation (N, %) | 3,022 (70.3%) |
| Length of Ventilation Episode, hours (Median, IQR) | 66.0 (29.0-118.0) |

Abbreviations: IQR, interquartile range; PIM-3, Pediatric Index of Mortality 3

The final cohort was randomly split into development (n=3,052), evaluation (n=611) and testing (n=763) datasets. To assess the impact of historical data on prediction performance, we evaluated models with varying lengths of clinical history from 1 to 48 hours (Figure 2). The initial task of nowcasting extubation outcome demonstrated a consistent decline in performance with increasing historical context (AUROC decreasing from 0.770 to 0.760, AUPRC from 0.420 to 0.396) and exhibited wider confidence intervals. In contrast, the second task – forecasting of extubation readiness demonstrated initial improvement with increasing historical context up to 24 hours (AUROC from 0.845 to 0.850, AUPRC from 0.432 to 0.451), followed by a gradual decline in AUPRC, while maintaining stable AUROC. The model exhibited narrow confidence intervals throughout. These patterns indicate different optimal historical context requirements for the two prediction tasks.

**Figure 2.**
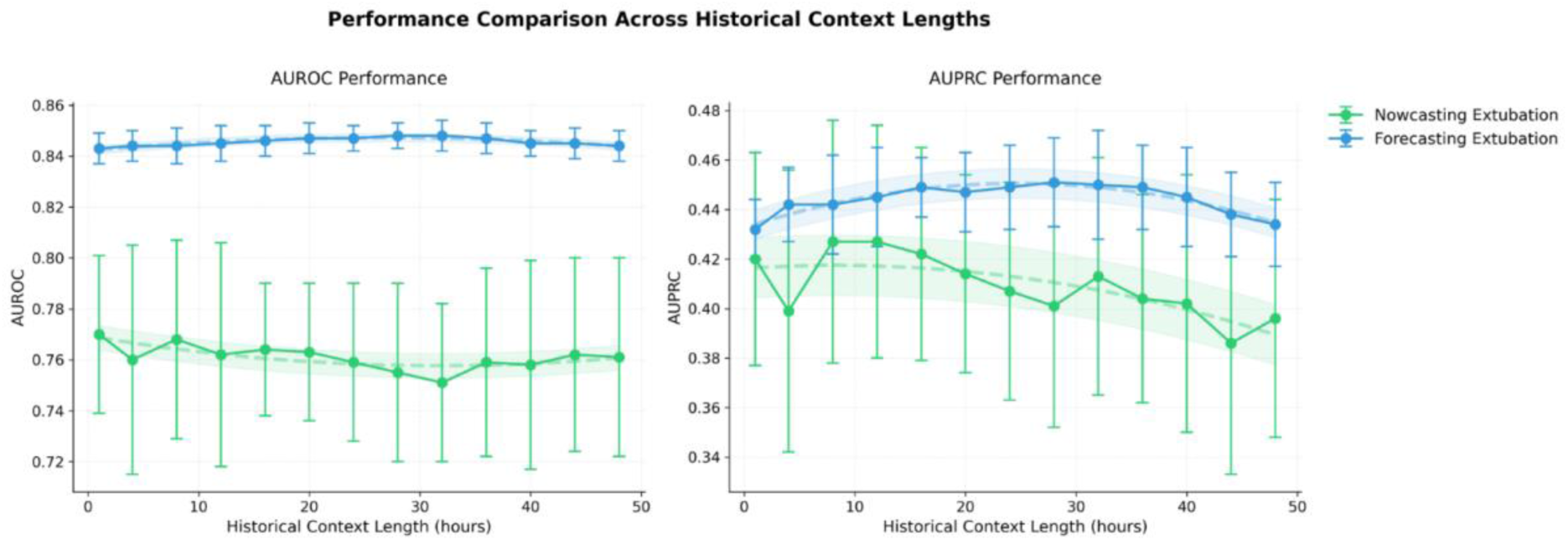
Model Performance Across Different Historical Context Lengths. Error bars indicate standard deviation from five repeated experiments with different random data splits. The forecasting model (blue) maintains stable performance (AUROC ∼0.85) across different history lengths, while the nowcasting model (green) shows optimal performance with shorter histories (AUROC ∼0.77). Both models were evaluated using area under the receiver operating characteristic curve (AUROC) and area under the precision-recall curve (AUPRC) for historical windows ranging from 1 to 48 hours.

The temporal evolution of risk scores preceding extubation exhibited distinct patterns between successful and failed cases as illustrated in Figure 3. For successful extubations, both the nowcasting risk score (green solid line) and the forecasting risk score (blue solid line) demonstrated a descending trajectory. Notably, the forecasting risk score accurately intersected the 0.5 decision threshold at the intended 12-hour planning window, indicating optimal timing prediction. In contrast, the nowcasting risk score incorrectly traversed this boundary earlier, at approximately 70 hours pre-extubation, potentially signalling extubation readiness prematurely. While showing some decline, cases of failed extubation (dashed lines), maintained consistently elevated risk scores in both tasks and displayed less improvement approaching extubation. The discrepancy between successful and failed cases was particularly evident in the forecasting risk assessment, with separation increasing as extubation approached, whereas the nowcasting risk scores showed more overlap and earlier boundary crossing than clinically appropriate.

**Figure 3.**
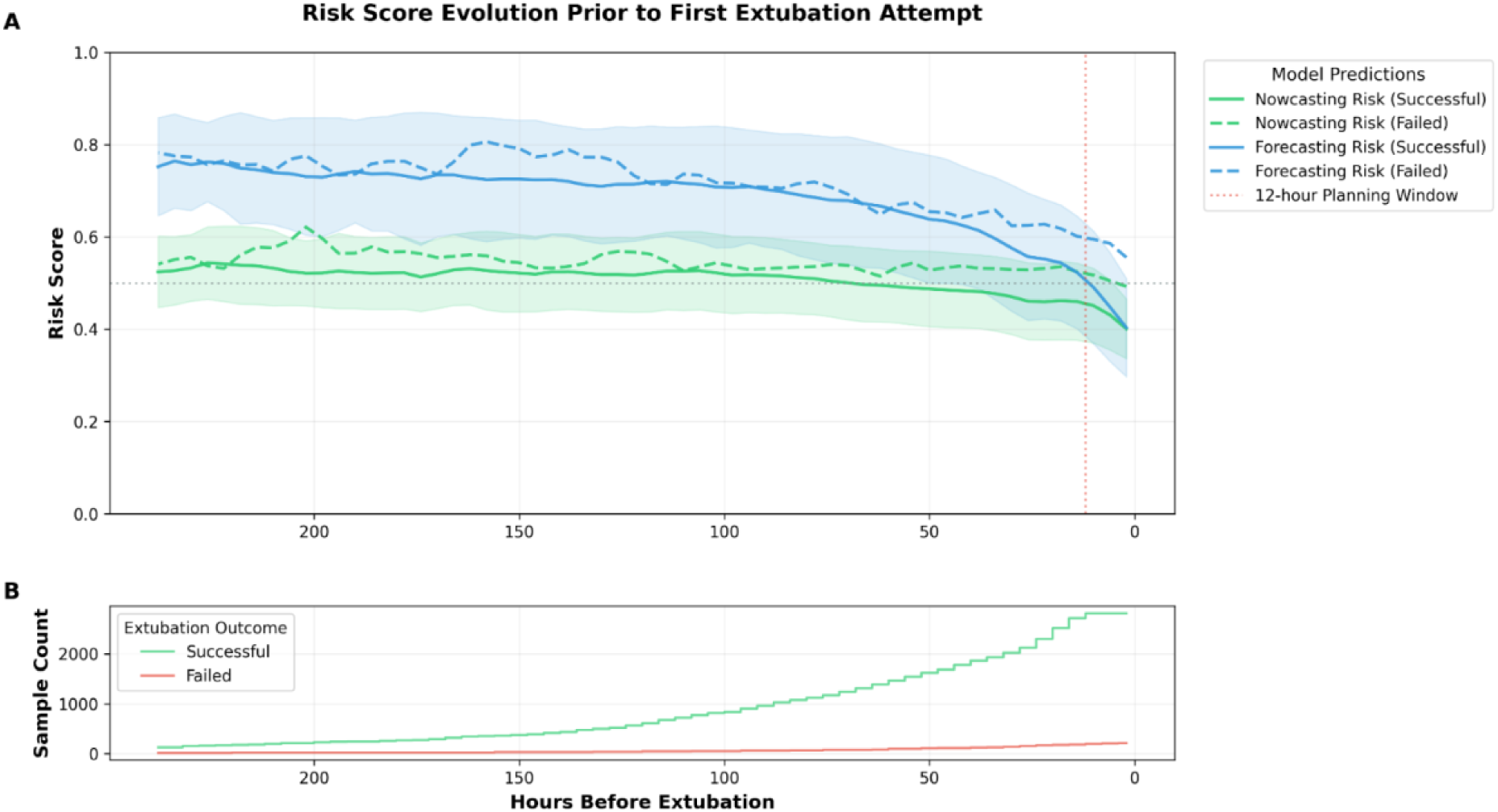
Risk Score Evolution and Sample Distribution Prior to Extubation. Panel A shows mean risk scores predicted by nowcasting (green) and forecasting (blue) models for successful (solid lines) and failed (dashed lines) extubation attempts over time. Shaded areas indicate 95% confidence intervals, with the vertical red dotted line marking the 12-hour planning window. Panel B presents sample count over time showing successful (green) and failed (red) extubations, with increasing data availability approaching the extubation event.

Feature importance analysis revealed distinct patterns between the two tasks (Figure 4). The nowcasting extubation model relied mostly on ventilation parameters, with the level of PS (in cmH_2_O) showing maximum importance, followed by ventilation mode and peak inspiratory pressure. In contrast, the forecasting extubation model demonstrated more distributed importance across multiple clinical domains, showing steroid use as the most important feature, with consistently high importance across medication interventions, physiological parameters, and demographic characteristics. Notably, while ventilation parameters dominated immediate prediction, their relative importance was substantially lower in forecasting task, for example, patient characteristics like sex had a relative importance of 0.86 and diagnostic group had a relative importance of 0.75 and therefore both gained prominence.

**Figure 4.**
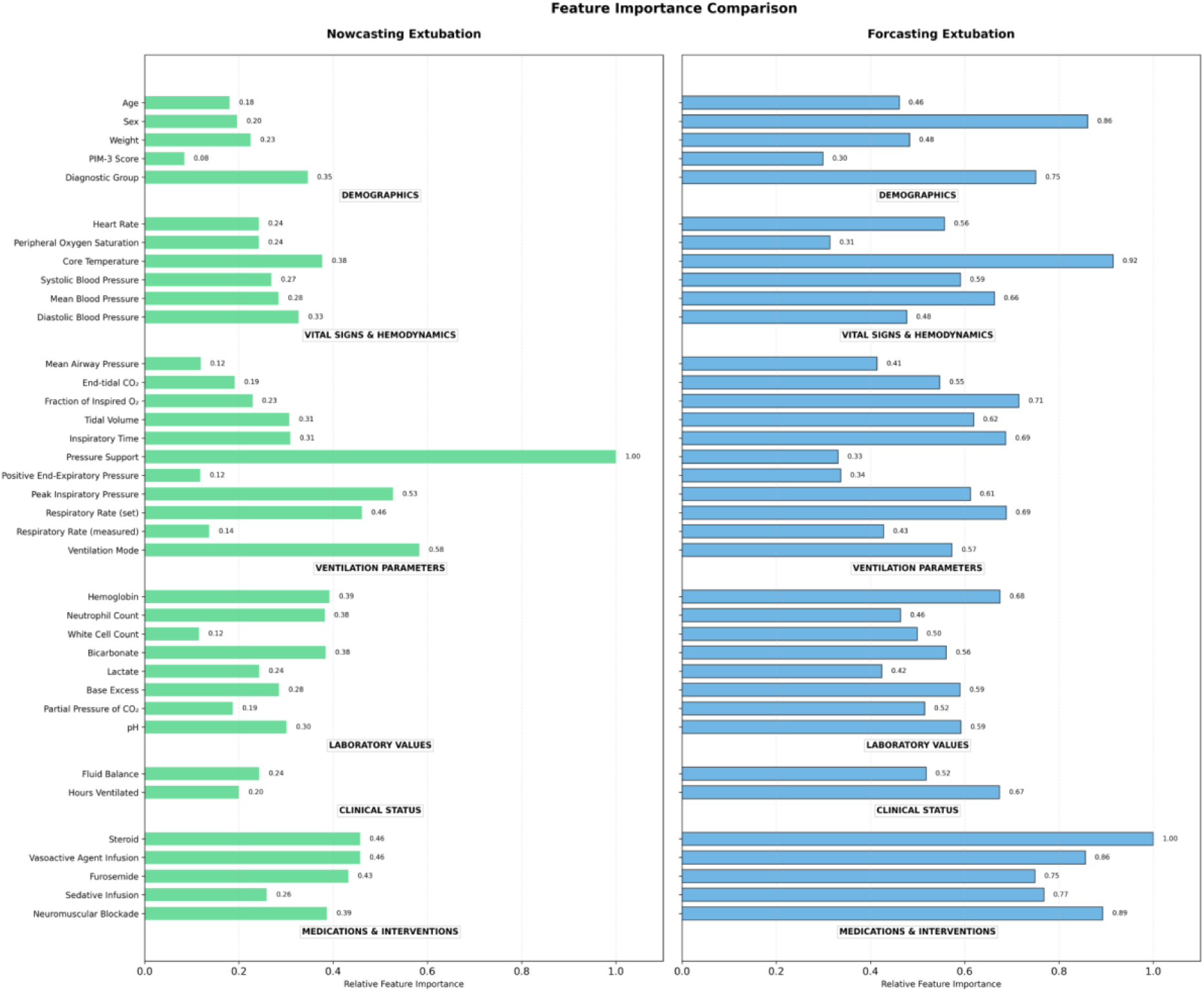
Feature Importance Comparison Between Prediction Models. Clinical features are grouped into six categories (demographics, vital signs and hemodynamics, ventilation parameters, laboratory values, clinical status, and medications and interventions), with their relative importance normalized to maximum values shown for both nowcasting (left, green) and forecasting (right, blue) models. The nowcasting model relied heavily on ventilation parameters, especially pressure support, while the forecasting model exhibited more distributed feature importance across categories, with strong influences from medications and patient characteristics.

## Discussion

In this large cohort study of 3,815 pediatric mechanical ventilation episodes, we developed and validated two complementary machine learning approaches for extubation decision support. The cohort’s characteristics reflected a diverse PICU population, with a broad range of ages (median 8.0 months), cardiac and respiratory conditions, and an extubation failure rate of 8.3% - comparable to previous reports from general and cardiac PICUs across the United Kingdom and in the United States.^28,35^

Our findings target a critical challenge in pediatric mechanical ventilation: balancing the risks of premature extubation against prolonged ventilation. While current practice relies heavily on point-in-time clinical assessment, our dual-model approach provides continuous and objective support for extubation decision-making. The nowcasting extubation model helps validate clinical judgment at the point of planned extubation, while the forecasting extubation model enables systematic monitoring of readiness, potentially helping identify optimal extubation windows that might otherwise be missed in routine care.

The differential temporal requirements of our two prediction tasks reveal important insights about the nature of extubation readiness assessment. While nowcasting extubation prediction performed best with recent clinical data, showing declining performance with extended history, the forecasting extubation prediction required longer observation periods (up to 24 hours) to achieve optimal performance. This pattern aligns with clinical intuition: immediate extubation success depends primarily on current physiological readiness, reflected in ventilation parameters and immediate clinical status. In contrast, identifying future optimal extubation timing requires consideration of broader clinical trajectory, including treatment response and overall stability. This is particularly relevant in the pediatric population, where rapid physiological changes are common and the traditional ‘snapshot’ approach to extubation assessment may miss important temporal trends.^36^

Analyzing feature importance across both models sheds light on the key factors influencing clinical decision-making in pediatric ventilation. For nowcasting extubation, the dominance of PS values and ventilation mode validates the clinical focus on respiratory independence parameters at extubation readiness assessment.^37^ For forecasting extubation, the high importance of steroid use and other medications, alongside patient characteristics, reinforces that successful extubation depends on broader clinical management and patient-specific factors. These findings suggest that while current bedside assessment appropriately focuses on ventilation parameters for immediate decisions, the identification of optimal extubation timing should additionally benefit from systematically considering medication response and patient factors. This is particularly relevant in the pediatric population, where recovery patterns vary significantly with age and diagnosis.

The literature on machine learning approaches for pediatric extubation prediction remains limited. The only comparable study, published in 2023, focused on low-birth-weight neonates (n=1,348) with an AUROC of 0.82 for the extubation failure prediction task.^16^ While they similarly identified ventilatory parameters as key predictors, our analysis of 3,815 episodes across a broader pediatric population provides more comprehensive insights for general PICU settings. Furthermore, our dual-model approach offers unique perspectives on the temporal nature of extubation readiness, suggesting a two-tiered strategy for clinical implementation: using longer-term trajectories to identify optimal extubation windows, followed by focused assessment of immediate physiological parameters to minimize failure risk. This strategy aligns with consensus that extubation success depends not only on meeting specific physiological criteria but also on the stability and direction of the patient’s overall clinical course.^8,37^

Several limitations of our study warrant discussion. The retrospective nature of our dataset required developing a decision tree to classify ventilation status, as no formal hourly classification existed. While we aimed to systematically extract variables, inconsistency in coding for manually entered data (e.g. medications) may have led to under-counting, potentially introducing misclassification despite careful validation. Our feature importance analysis, using absolute values across the temporal dimension to prevent offsetting effects, limited our ability to determine whether features had positive or negative impacts at different timepoints. Additionally, several potentially predictive factors could not be incorporated in the model, such as resolution of underlying illness, need for other procedures, consciousness level, and unit acuity.

Regarding generalizability, our focus on first extubation and traditional ventilation modes, while enabling analysis of common clinical scenarios, may not extend to more complex cases requiring volume control, high-frequency oscillatory ventilation, or those with tracheostomy in-situ. Centre-specific practices regarding ventilation and extubation may also limit external validity. The exclusion of patients who died within 48 hours post-extubation, while necessary to avoid confounding from planned terminal extubations, may have omitted useful information. Finally, while our models demonstrate robust discrimination in retrospective testing, prospective validation in real-world clinical settings is needed to assess their impact on patient outcomes.

## Conclusion

In this large multicenter study, we developed and validated two complementary machine learning approaches for predicting extubation readiness in critically ill children. Our nowcasting extubation outcome prediction model provides point-of-care decision support for extubation attempts, while the forecasting extubation prediction model enables continuous monitoring of patient readiness. The distinct patterns observed between these models suggest a practical approach to extubation decision-making: continuous monitoring to identify optimal extubation windows, supported by focused physiological assessment at the time of planned extubation. While prospective validation is needed, this novel dual-model strategy offers a structured framework that balances the risks of prolonged ventilation against premature extubation in the pediatric intensive care setting.

## Acknowledgements

Author Contributions:

Y.L. led the study design, performed data analysis, developed the machine learning models, and wrote the initial manuscript draft. S.C.A. and E.C.A. provided clinical expertise in data interpretation and contributed substantially to manuscript writing. J.O. provided technical support and methodology validation. T.C.A., A.A., and S.R. facilitated data collection at their respective clinical sites and contributed clinical expertise in data interpretation. A.A.F. and P.R. conceptualized the study, secured funding, provided access to the data, and supervised the project. All authors critically reviewed and approved the final manuscript.

## Data Availability

The datasets generated and/or analyzed during the current study are not publicly available due to patient privacy regulations and institutional policies but are available from the corresponding author on reasonable request.

## Code Availability

The underlying code for this study is not publicly available but may be made available to qualified researchers upon reasonable request from the corresponding author.

## Patient and public involvement

No patient or public involvement was undertaken in this research.

## References

1. PICANet. PICANet State of the Nation Report 2022, <https://www.picanet.org.uk/annual-reporting-and-publications/> (2022).

2 Liu, Y. et al. Characteristics and Risk Factors of Children Requiring Prolonged Mechanical Ventilation vs. Non-prolonged Mechanical Ventilation in the PICU: A Prospective Single-Center Study. Frontiers in Pediatrics 10 (2022).

3 Loss, S. et al. The reality of patients requiring prolonged mechanical ventilation: A multicenter study. Revista Brasileira de Terapia Intensiva 27 (2015). 10.5935/0103-507X.20150006

4 Khemani, R. G. et al. Risk Factors for Pediatric Extubation Failure: The Importance of Respiratory Muscle Strength*. Crit Care Med 45 (2017).

5 Thille, A. W., Richard, J.-C. M. & Brochard, L. The Decision to Extubate in the Intensive Care Unit. American Journal of Respiratory and Critical Care Medicine 187, 1294–1302 (2013). 10.1164/rccm.201208-1523CI

6 Loberger, J. M., Manchikalapati, A., Borasino, S. & Prabhakaran, P. Prevalence, Risk Factors, and Outcomes of Airway Versus Non-Airway Pediatric Extubation Failure. Respiratory Care 68, 374 (2023). 10.4187/respcare.10341

7 Miura, S., Butt, W., Thompson, J. & Namachivayam, S. P. Recurrent Extubation Failure Following Neonatal Cardiac Surgery Is Associated with Increased Mortality. Pediatric Cardiology 42, 1149–1156 (2021). 10.1007/s00246-021-02593-2

8 Abu-Sultaneh, S. et al. Executive Summary: International Clinical Practice Guidelines for Pediatric Ventilator Liberation, A Pediatric Acute Lung Injury and Sepsis Investigators (PALISI) Network Document. American Journal of Respiratory and Critical Care Medicine 207, 17–28 (2022). 10.1164/rccm.202204-0795SO

9 Kim, Y. et al. Early prediction of need for invasive mechanical ventilation in the neonatal intensive care unit using artificial intelligence and electronic health records: a clinical study. BMC Pediatrics 23, 525 (2023). 10.1186/s12887-023-04350-1

10 Huang, K.-Y. et al. Developing a machine-learning model for real-time prediction of successful extubation in mechanically ventilated patients using time-series ventilator-derived parameters. Frontiers in Medicine 10 (2023). 10.3389/fmed.2023.1167445

11 Johnston, C., de Carvalho, W. B., Piva, J., Garcia, P. C. R. & Fonseca, M. C. Risk factors for extubation failure in infants with severe acute bronchiolitis. Respiratory care 55, 328–333 (2010).

12 Laham, J. L., Breheny, P. J. & Rush, A. Do Clinical Parameters Predict First Planned Extubation Outcome in the Pediatric Intensive Care Unit? Journal of Intensive Care Medicine 30, 89–96 (2013). 10.1177/0885066613494338

13 Iyer, N. P. et al. Association of Extubation Failure Rates With High-Flow Nasal Cannula, Continuous Positive Airway Pressure, and Bilevel Positive Airway Pressure vs Conventional Oxygen Therapy in Infants and Young Children: A Systematic Review and Network Meta-Analysis. JAMA Pediatrics 177, 774–781 (2023). 10.1001/jamapediatrics.2023.1478

14 Wang, Z. et al. Developing an explainable machine learning model to predict the mechanical ventilation duration of patients with ARDS in intensive care units. Heart & Lung: The Journal of Cardiopulmonary and Acute Care 58, 74–81 (2023). 10.1016/j.hrtlng.2022.11.005

15 Sayed, M., Riaño, D. & Villar, J. Predicting Duration of Mechanical Ventilation in Acute Respiratory Distress Syndrome Using Supervised Machine Learning. Journal of Clinical Medicine 10 (2021).

16 Natarajan, A. et al. Prediction of extubation failure among low birthweight neonates using machine learning. Journal of Perinatology 43, 209–214 (2023). 10.1038/s41372-022-01591-3

17 Tao, Y., Ding, X. & Guo, W.-l. Using machine-learning models to predict extubation failure in neonates with bronchopulmonary dysplasia. BMC Pulmonary Medicine 24, 308 (2024). 10.1186/s12890-024-03133-3

18 Wu, J. T., Leung, K. & Leung, G. M. Nowcasting and forecasting the potential domestic and international spread of the 2019-nCoV outbreak originating in Wuhan, China: a modelling study. The Lancet 395, 689–697 (2020). 10.1016/S0140-6736(20)30260-9

19 Sarti, A. J. et al. Feasibility of implementing ‘Extubation Advisor’, a clinical decision support tool to improve extubation decision-making in the ICU: a mixed-methods observational study. BMJ Open 11, e045674 (2021). 10.1136/bmjopen-2020-045674

20 Collins, G. S. et al. TRIPOD+AI statement: updated guidance for reporting clinical prediction models that use regression or machine learning methods. BMJ 385, e078378 (2024). 10.1136/bmj-2023-078378

21 Neto, J., Casimiro, H. J. & Reis-Pina, P. Palliative Extubation in Pediatric Patients in the Intensive Care Unit and at Home: A Scoping Review. International Journal of Pediatrics 2023, 6697347 (2023). 10.1155/2023/6697347

22 Kawaguchi, A. et al. Longventkids Study: A Prospective Cohort Study on Prolonged Mechanical Ventilated Children. (2024).

23 Keselman, H. & Rogan, J. The Tukey multiple comparison test: 1953-1976. Psychological Bulletin 84, 1050–1056 (1977). 10.1037/0033-2909.84.5.1050

24 Vedrenne-Cloquet, M. et al. Pleural and transpulmonary pressures to tailor protective ventilation in children. Thorax 78, 97 (2023). 10.1136/thorax-2021-218538

25. Hug, C. W. *Detecting hazardous intensive care patient episodes using real-time mortality models* PhD thesis, Massachusetts Institute of Technology, (2009).

26 Tutz, G. & Ramzan, S. Improved methods for the imputation of missing data by nearest neighbor methods. Computational Statistics & Data Analysis 90, 84–99 (2015). 10.1016/j.csda.2015.04.009

27 Mastropietro, C. W. et al. Extubation Failure after Neonatal Cardiac Surgery: A Multicenter Analysis. J Pediatr 182, 190–196.e194 (2017). 10.1016/j.jpeds.2016.12.028

28 Ramnarayan, P. et al. Effect of High-Flow Nasal Cannula Therapy vs Continuous Positive Airway Pressure Following Extubation on Liberation From Respiratory Support in Critically Ill Children: A Randomized Clinical Trial. JAMA 327, 1555–1565 (2022). 10.1001/jama.2022.3367

29 Baisch, S. D., Wheeler, W. B., Kurachek, S. C. & Cornfield, D. N. Extubation failure in pediatric intensive care incidence and outcomes. Pediatric Critical Care Medicine 6 (2005).

30 Newth, C. J. L. et al. Weaning and extubation readiness in pediatric patients*. Pediatric Critical Care Medicine 10 (2009).

31 Cho, K. et al. Learning Phrase Representations using RNN Encoder-Decoder for Statistical Machine Translation. (2014). 10.3115/v1/D14-1179

32 Lin, T.-Y., Goyal, P., Girshick, R., He, K. & Dollar, P. Focal Loss for Dense Object Detection. IEEE Transactions on Pattern Analysis and Machine Intelligence PP, 1–1 (2018). 10.1109/TPAMI.2018.2858826

33. Sundararajan, M., Taly, A. & Yan, Q. in Proceedings of the 34th International Conference on Machine Learning Vol. 70 (eds Precup Doina & Teh Yee Whye) 3319--3328 (PMLR, Proceedings of Machine Learning Research, 2017).

34. Ismail, A., Gunady, M., Corrada Bravo, H. & Feizi, S. Benchmarking Deep Learning Interpretability in Time Series Predictions. (2020).

35. Gaies, M., et al. Clinical Epidemiology of Extubation Failure in the Pediatric Cardiac ICU: A Report From the Pediatric Cardiac Critical Care Consortium*. Pediatric Critical Care Medicine 16 (2015).

36 Carter, M. J. et al. Evaluation of Phoenix Sepsis Score Criteria: Exploratory Analysis of Characteristics and Outcomes in an Emergency Transport PICU Cohort From the United Kingdom, 2014–2016. Pediatric Critical Care Medicine (2025).

37 Torrini, F. et al. Prediction of extubation outcome in critically ill patients: a systematic review and meta-analysis. Critical Care 25, 391 (2021). 10.1186/s13054-021-03802-3

