## Supplemental Information for "Machine Learning Models for Dynamic Assessment of Extubation Readiness in Pediatric Critical Care"

This supplemental material has been provided by the authors to give readers additional information about their work.

Contents:

eTable 1. Features of Interest Used in Prediction Models

eTable 2. Class-wise Comparison of Clinical Characteristics in Two Prediction Tasks

eFigure 1. Study Flow Diagram of Patient Selection and Data Split

#### eTable 1. Features of Interest Used in Prediction Models

| **Demographics and outcomes** | **Hourly vital signs and ventilator data** |
|---|---|
| Age | Heart rate (bpm) |
| Sex | Systolic blood pressure (mm Hg) |
| Weight (kg) | Mean blood pressure (mm Hg) |
| Diagnostic group | Diastolic blood pressure (mm Hg) |
| PIM-3 score | Core temperature (C) |
| **Blood gases and laboratory tests** | Mean airway pressure (cm H2O) |
| pH | Measured respiratory rate |
| pCO2 (kPa) | End tidal CO2 (kPa) |
| Base excess (mmol/L) | Peripheral oxygen saturation |
| Lactate (mmol/L) | Tidal volume (ml/kg) |
| Bicarbonate (mmol/L) | Hours on ventilator |
| White cell count | Ventilator mode† |
| Neutrophil count | Fraction of inspired oxygen |
| Haemoglobin (g/L) | Peak inspiratory pressure |
| **Medications (yes/no)** | Positive end-expiratory pressure |
| Sedative infusion | Pressure Support (cm H2O) |
| Neuromuscular blockade | Set respiratory rate |
| Vasoactive agent infusion | Inspiratory time (seconds) |
| Furosemide | Net fluid balance (ml) |
| Corticosteroids | |

† Two ventilation modes were included in the analysis: (1) Pressure control (PC): Pressure control-biphasic positive airway pressure (PC-BIPAP), which aligns with pressure control intermittent mandatory ventilation with set-point targeting scheme (PC-IMVs) in international ventilation taxonomy; (2) Pressure Support (PS): Spontaneous continuous positive airway pressure with pressure support (SPN-CPAP PS), which aligns with pressure control continuous spontaneous ventilation (PC-CSV) in international ventilation taxonomy.

### eTable 2. Class-wise Comparison of Clinical Characteristics in Two Prediction Tasks

| Features | Extubation Success (n=3500) | Extubation Failure (n=315) | p-value | Effect Size | Not Ready-to-Extubate (n=248156) | Ready-to-Extubate (n=42000) | p-value | Effect Size |
|---|---|---|---|---|---|---|---|---|
| Ventilator Hours | 65.00 (30.00 - 113.00) | 55.00 (25.50 - 108.00) | 0.024 | -0.076 | 137.00 (88.00 - 233.00) | 65.00 (30.00 - 113.00) | <0.001 | -0.527 |
| Neuromuscular blockade | 8 (0.2%) | 22 (7.0%) | <0.001 | N/A | 18053 (7.3%) | 476 (1.1%) | <0.001 | N/A |
| Sedative infusion | 1252 (35.8%) | 152 (48.3%) | <0.001 | N/A | 119380 (48.1%) | 20859 (49.7%) | <0.001 | N/A |
| Furosemide | 395 (11.3%) | 39 (12.4%) | 0.621 | N/A | 30021 (12.1%) | 5037 (12.0%) | 0.548 | N/A |
| Vasoactive agent infusion | 836 (23.9%) | 96 (30.5%) | 0.011 | N/A | 59108 (23.8%) | 10358 (24.7%) | <0.001 | N/A |
| Corticosteroids | 247 (7.1%) | 15 (4.8%) | 0.154 | N/A | 1353 (0.5%) | 1251 (3.0%) | <0.001 | N/A |
| Ventilator mode (PC) | 1258 (35.9%) | 206 (65.4%) | <0.001 | N/A | 233311 (94.0%) | 27672 (65.9%) | <0.001 | N/A |
| Ventilator mode (PS) | 2242 (64.1%) | 109 (34.6%) | <0.001 | N/A | 14845 (6.0%) | 14328 (34.1%) | <0.001 | N/A |
| Mean airway pressure (cm H2O) | 7.00 (6.00 - 8.00) | 8.00 (7.00 - 9.00) | <0.001 | 0.375 | 9.00 (8.00 - 11.00) | 7.00 (7.00 - 8.00) | <0.001 | -0.535 |
| Measured respiratory rate | 28.00 (21.00 - 38.00) | 29.00 (24.00 - 37.00) | 0.078 | 0.060 | 26.00 (22.00 - 33.00) | 26.00 (20.00 - 35.00) | 0.242 | 0.004 |
| End tidal CO2 (kPa) | 5.20 (4.50 - 5.90) | 5.10 (4.30 - 5.80) | 0.005 | -0.095 | 5.20 (4.48 - 6.00) | 5.20 (4.60 - 5.90) | 0.001 | 0.010 |
| Core temperature (C) | 37.20 (36.80 - 37.50) | 37.00 (36.55 - 37.50) | <0.001 | -0.153 | 37.00 (36.50 - 37.40) | 37.20 (36.80 - 37.50) | <0.001 | 0.150 |
| Heart rate (bpm) | 125.00 (108.00 - 140.00) | 132.00 (112.00 - 148.00) | <0.001 | 0.154 | 127.00 (110.00 - 143.00) | 123.00 (107.00 - 138.00) | <0.001 | -0.101 |
| Peripheral oxygen saturation | 97.00 (95.00 - 99.00) | 97.00 (93.00 - 99.00) | 0.122 | -0.052 | 97.00 (94.00 - 99.00) | 97.00 (95.00 - 99.00) | <0.001 | 0.017 |
| Systolic blood pressure (mm Hg) | 90.00 (77.00 - 104.00) | 82.00 (70.00 - 98.50) | <0.001 | -0.195 | 81.00 (69.00 - 95.00) | 86.00 (74.00 - 99.00) | <0.001 | 0.127 |
| Mean blood pressure (mm Hg) | 63.00 (54.00 - 73.00) | 57.00 (49.00 - 68.00) | <0.001 | -0.182 | 56.00 (48.00 - 65.00) | 58.00 (50.00 - 68.00) | <0.001 | 0.127 |
| Diastolic blood pressure (mm Hg) | 49.00 (41.00 - 58.00) | 44.00 (38.00 - 53.50) | <0.001 | -0.164 | 43.00 (36.00 - 51.00) | 45.00 (38.00 - 54.00) | <0.001 | 0.119 |
| pH | 7.41 (7.37 - 7.45) | 7.39 (7.34 - 7.44) | <0.001 | -0.166 | 7.39 (7.35 - 7.44) | 7.41 (7.36 - 7.45) | <0.001 | 0.109 |
| PCO2 (kPa) | 5.89 (5.20 - 6.70) | 5.92 (5.18 - 6.78) | 0.834 | -0.007 | 6.14 (5.30 - 7.29) | 5.90 (5.20 - 6.79) | <0.001 | -0.114 |
| Base excess (mmol/L) | 3.20 (-0.40 - 7.50) | 1.60 (-2.30 - 5.85) | <0.001 | -0.156 | 3.10 (-0.80 - 8.30) | 2.90 (-1.00 - 7.60) | <0.001 | -0.023 |
| Lactate (mmol/L) | 1.00 (0.70 - 1.30) | 1.10 (0.80 - 1.60) | <0.001 | 0.165 | 1.00 (0.79 - 1.40) | 1.00 (0.70 - 1.30) | <0.001 | -0.047 |
| Bicarbonate (mmol/L) | 27.00 (23.90 - 31.30) | 25.60 (22.20 - 29.50) | <0.001 | -0.167 | 27.30 (23.90 - 32.20) | 26.80 (23.60 - 31.30) | <0.001 | -0.047 |
| White cell count | 9.66 (7.20 - 12.80) | 10.10 (7.60 - 13.60) | 0.066 | 0.063 | 9.50 (6.90 - 13.10) | 9.70 (7.20 - 12.80) | 0.001 | 0.010 |
| Neutrophil count | 5.70 (3.60 - 8.41) | 6.19 (3.70 - 9.06) | 0.074 | 0.061 | 5.50 (3.30 - 8.60) | 5.70 (3.60 - 8.50) | <0.001 | 0.018 |
| Haemoglobin (g/L) | 107.00 (94.00 - 122.00) | 111.00 (96.00 - 129.00) | 0.003 | 0.100 | 104.00 (92.00 - 122.00) | 107.00 (94.00 - 123.00) | <0.001 | 0.057 |
| Net fluid balance (ml) | -0.80 (-19.00 - 10.00) | 2.30 (-13.95 - 10.75) | 0.013 | 0.084 | 3.00 (-11.00 - 17.00) | 0.70 (-15.00 - 12.80) | <0.001 | -0.083 |
| Fraction of inspired oxygen | 25.00 (21.00 - 30.00) | 30.00 (25.00 - 40.00) | <0.001 | 0.226 | 30.00 (25.00 - 40.00) | 25.00 (21.00 - 31.00) | <0.001 | -0.268 |
| Peak inspiratory pressure | 0.00 (0.00 - 15.00) | 15.00 (0.00 - 19.00) | <0.001 | 0.406 | 18.00 (15.00 - 21.00) | 15.00 (0.00 - 16.00) | <0.001 | -0.561 |
| Set respiratory rate | 0.00 (0.00 - 5.00) | 10.00 (0.00 - 22.00) | <0.001 | 0.431 | 20.00 (14.00 - 25.00) | 5.00 (0.00 - 15.00) | <0.001 | -0.555 |
| Positive end-expiratory pressure | 5.00 (5.00 - 5.00) | 5.00 (5.00 - 5.00) | <0.001 | 0.129 | 5.00 (5.00 - 6.00) | 5.00 (5.00 - 5.00) | <0.001 | -0.268 |
| Inspiratory time (seconds) | 0.00 (0.00 - 0.80) | 0.70 (0.00 - 0.85) | <0.001 | 0.263 | 0.80 (0.70 - 1.00) | 0.70 (0.00 - 0.89) | <0.001 | -0.243 |
| Pressure support (cm H2O) | 5.00 (0.00 - 8.00) | 0.00 (0.00 - 5.00) | <0.001 | -0.274 | 0.00 (0.00 - 0.00) | 0.00 (0.00 - 5.00) | <0.001 | 0.281 |
| Tidal volume (ml/kg) | 7.22 (5.45 - 9.33) | 6.96 (5.13 - 8.93) | 0.112 | -0.054 | 7.18 (5.61 - 8.86) | 7.29 (5.62 - 9.25) | <0.001 | 0.036 |

For the nowcasting extubation outcome prediction task (left columns), characteristics were measured in the hour immediately preceding extubation attempts (n=3,815). For the forecasting readiness prediction task (right columns), characteristics were measured at hourly intervals throughout the ventilation period (n=290,156). For time-varying measurements, values represent the last measurement in the observation window. Continuous variables are presented as median (IQR) and categorical variables as count (percentage). Effect sizes for continuous variables were calculated using Cohen's d, while N/A indicates categorical variables where effect size is not applicable. While the prediction models were trained with varying historical window sizes (1-48 hours) as a hyperparameter, for the purpose of this baseline comparison, we look at only the last measurements to ensure consistent comparison across all cases.

**eFigure 1. Study Flow Diagram of Patient Selection and Data Split**

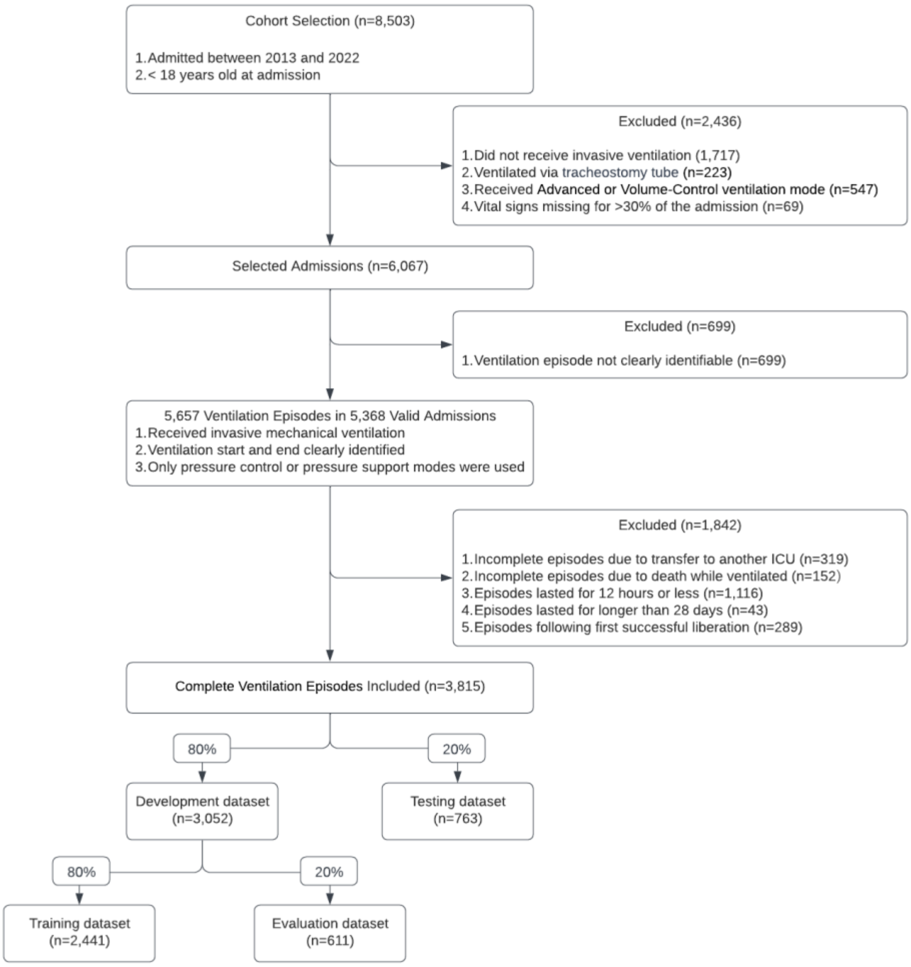


Flow diagram showing patient selection, exclusion criteria, and data partitioning strategy for model development. From 8,503 initial admissions, 3,815 complete ventilation episodes were included after applying exclusion criteria. The final cohort was partitioned into development and testing sets, with further subdivision of the development set for model training and evaluation.
